# Mortality associated with different influenza subtypes in France between 2015-2019

**DOI:** 10.1101/2022.11.21.22282612

**Authors:** Edward Goldstein

## Abstract

**Background:** High levels of excess mortality during periods of active influenza circulation in France were observed in the years preceding the COVID-19 pandemic. Some of the factors that affect the rates of influenza associated mortality are influenza vaccination coverage levels in different population groups and practices for testing for influenza and related use of antiviral medications for various illness episodes (including pneumonia hospitalizations) during periods of active influenza circulation in the community.

**Methods:** Data on sentinel ILI surveillance and sentinel virological surveillance in France were combined in a framework of a previously developed regression model to estimate the number of deaths associated with the circulation of the major influenza subtypes (A/H3N2, A/H1N1, B/Yamagata and B/Victoria) in France between 2015-2019.

**Results:** Between week 3, 2015 and week 2, 2020, there were on average 15403 (95% CI (12591,18229)) annual influenza-associated deaths, of which 60.3% (49.9%,71.9%) were associated with influenza A/H3N2, and 29.5% (13.3%,45.5%) were associated with influenza B/Yamagata. During weeks when levels of ILI consultation in mainland France were above 50 per 100,000 persons, 7.9% (6.5%,9.4%) of all deaths in France were influenza-associated.

**Conclusions:** High rates of influenza-associated mortality in France prior to the COVID-19 pandemic suggest that boosting influenza vaccination coverage in different population groups and testing for influenza in respiratory illness episodes (including pneumonia hospitalizations) during periods of active influenza (particularly influenza A/H3N2) circulation in combination with the use of antiviral medications is needed to mitigate the impact of influenza epidemics.

## Introduction

Prior to the COVID-19 pandemic, annual influenza epidemics resulted in high rates of influenza-associated mortality in different countries [1-5] (e.g. an estimated average annual rate of over 16 deaths per 100,000 individuals in the EU countries for the 2012-2013 through the 2017-2018 influenza seasons [5]). Only a fraction of influenza-associated mortality has respiratory diseases as an underlying cause of death [1,2,6], with an additional significant contribution of influenza infections to mortality for circulatory and other non-respiratory causes [1,6,7]. Despite the high rates of influenza-associated mortality, frequency of testing for influenza and related use of antiviral medications may be relatively low for various illness episodes (including pneumonia hospitalizations) during periods of active influenza circulation in the community. A US study found that frequency of testing for influenza in cases of community-acquired pneumonia (CAP) in the US had increased significantly between 2010-2015, whereas oseltamivir use on the first day of hospitalization in influenza-positive cases was associated with a relative risk of 0.75; 95% CI, 0.59-0.96 for 14-day in-hospital mortality [8]. Among the different influenza subtypes, influenza A/H3N2 was found to carry the greatest relative contribution to influenza-associated mortality [1-3,5]. The older age distribution for cases of severe outcomes associated with influenza A/H3N2 infections compared to influenza A/H1N1/pdm infections [9] is related to the first influenza infection experienced by individuals in different age groups [10]. For cases of influenza-associated hospitalization, influenza A/H1N1pdm carries a much higher risk of pneumonia compared to influenza A/H3N2 [9], though A/H1N1pdm hospitalizations have a significantly younger age distribution compared to A/H3N2 hospitalizations (Table 1 in ref. [9]). This suggests that rates of influenza-associated pneumonia in non-elderly individuals can be high during A/H1N1 epidemics. Distribution of cases for influenza B/Yamagata is significantly older compared to influenza B/Victoria [11], and epidemics associated with influenza B/Yamagata during select seasons were found to be associated with a significant influenza mortality burden [5], though levels of circulation of influenza B/Yamagata during the COVID-19 pandemic have been very low [12].

**Table 1:** Average annual number of deaths associated with different influenza subtypes in France, 2015-2019

| Influenza subtype | Average annual number of deaths | Percent of total influenza-associated deaths |
| --- | --- | --- |
| A/H1N1 | 641 (-2969,4239) | 4.1% (-19.9%,27.7%) |
| A/H3N2 | 9253 (7431,11059) | 60.3% (49.9%,71.9%) |
| B/Victoria | 959 (-294,2203) | 6.1% (-2%,13.8%) |
| B/Yamagata | 4551 (1925,7199) | 29.5% (13.3%,45.5%) |
| All influenza | 15403 (12591,18229) | 100% |

Rates of excess mortality associated with influenza epidemics vary between countries (e.g. Figure 1 in [13]), with that variability being related to differences in the age distribution for the population, differences in influenza vaccination coverage (including vaccination coverage in risk groups and vaccination coverage in groups that play the largest role in the spread on influenza in the community, particularly children [14]) and other factors. In France, in the years preceding the COVID-19 pandemic, high levels of excess mortality were observed during periods of active influenza circulation (e.g. Figure 8 in [15]). Influenza vaccination coverage levels in different population groups in France are moderate-to-low [16], and significantly lower than influenza vaccination coverage levels in the corresponding population groups in the US [17]. Additionally, testing for influenza during respiratory illness episodes and the related use of antiviral medications in the US has increased in recent years [5]. The aim of this study is to better characterize mortality associated with the circulation of different influenza subtypes in France to inform efforts on influenza vaccination in different population groups, and on testing for influenza in respiratory illness episodes (including pneumonia hospitalizations) during periods of active influenza circulation, and the related use of antiviral medications. Additionally, very high rates of influenza-associated mortality in individuals aged over 75y during certain influenza seasons in France [15] suggest that additional efforts are needed to better protect those populations during influenza epidemics, both in nursing homes and in the general community.

**Figure 1:**
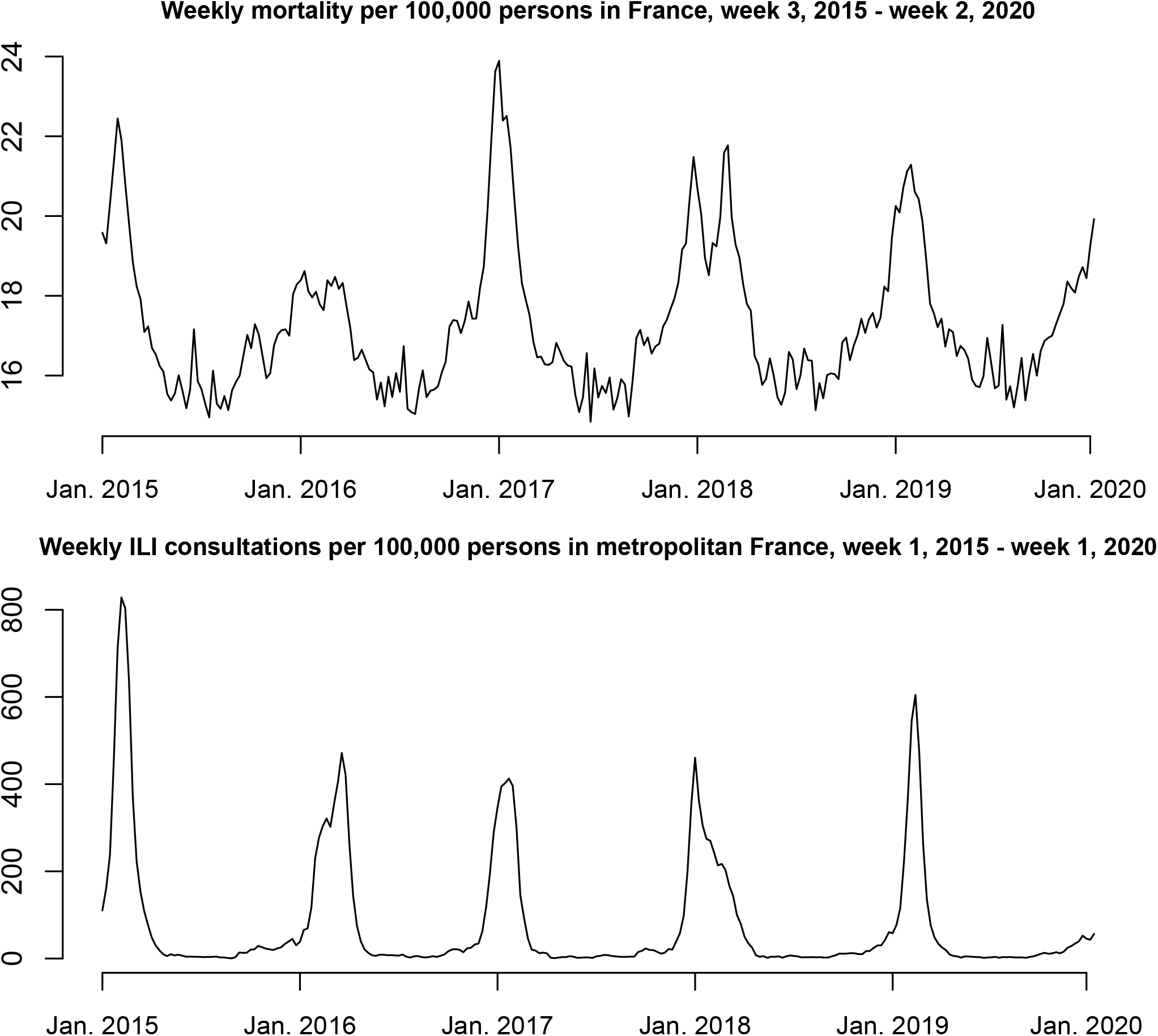
Weekly mortality per 100,000 persons in France, week 3, 2015 - week 2, 2020 and weekly rates of ILI consultations in Metropolitan France, week 2, 2015 - week 1, 2020

Excess mortality in France is known to be related to elevated rates of influenza-like illness (ILI) consultations [18,19] (see also Figure 1 in this paper). Additionally, the types of influenza strains circulating in France affect the rates of influenza-associated mortality. For example, the 2015-2016 epidemic in France was dominated by influenza A/H1N1 and B/Victoria (WHO FluNET viral surveillance data [20]) and had low levels of excess/influenza-associated mortality compared to the 2016-2017 epidemic, that was dominated by influenza A/H3N2 and had high levels of excess/influenza-associated mortality, particularly among individuals aged over 75y (Figure 8 in [15]), even though rates of ILI consultation were similar for those two seasons [19]. Lower levels of excess mortality associated with the circulation of influenza A/H1N1 and B/Victoria is related to the younger age distribution for cases of A/H1N1 and B/Victoria illness [9-11] mentioned in the previous paragraph. Another factor that affects the age distribution of influenza cases and the relation between rates of ILI medical consultations in the population and rates of influenza-associated mortality is the antigenic novelty of influenza strains, with antigenically novel strains (e.g. during the 2014-2015 A/H3N2 epidemic [21]) generally having a younger age distribution of cases of influenza infection. In our earlier work [1,6], we developed a method for combing data on syndromic surveillance with data on virologic surveillance to estimate rates of mortality associated with the major influenza subtypes in the US. Subsequently, this method was applied to the estimation of influenza-associated mortality in other countries [3-5], including the EU population [5]. Here, we apply this method to data on sentinel ILI surveillance in France [19], data on sentinel virological surveillance in France (available since 2015 [20]) and data on daily deaths in France since 2015 [22] to evaluate mortality associated with the major influenza subtypes in France between 2015-2019.

## Methods

### Data

Data on weekly numbers of ILI consultations in metropolitan France are available from the French sentinel surveillance [19]. Sentinel data on testing of respiratory specimens for the different influenza subtypes are available from WHO FluNET [20]. Data on the daily number of deaths in France starting 2015 are available from [22]. Data on population in France are available from [23].

### Statistical model

Not all ILI consultations in the sentinel data [19] correspond to influenza infections, and those that do, correspond to infection with different influenza subtypes. For each influenza subtype (e.g. A/H3N2), we define an indicator for the incidence of that subtype on week *t* (e.g. *A/H3N2(t*)) as

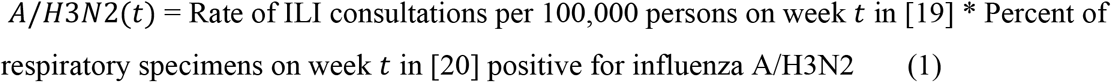

We note that during weeks of active influenza circulation (high levels of ILI in [19]) between 2015-2019, over 60% of respiratory specimens in [20] were positive for influenza (for at least one influenza subtype, Figure 2). To relate the incidence indicators for the major influenza subtypes to weekly levels of all-cause mortality per 100,000 persons in France [22], we note that age distribution of influenza cases changes with the appearance of antigenically novel influenza strains, and this changes the relation between rates of influenza-associated ILI (incidence indicators in eq. 1) and rates of influenza-associated mortality. The relevant antigenic changes for our study period were (a) the appearance of an antigenically/genetically novel A/H3N2 strain during the 2014-2015 influenza season [21]; the appearance of the novel influenza B/Yamagata strain during the 2017-2018 season [5]; the 2018-2019 A/H3N2 epidemic strains belonging to several clades and exhibiting different immunity profiles for different birth cohorts [24]. Correspondingly, for the model relating A/H3N2 circulation to associated mortality, we split the A/H3N2 incidence indicator into three: *A/H3N2*_1_, equaling the A/H3N2 incidence indicator for the period from Jan. 2015 to Sep. 2015, and equaling to 0 for later weeks; *A/H3N2*_2_, equaling the A/H3N2 incidence indicator for the period from Oct. 2015 to Sep. 2018, and equaling to 0 for other weeks, and *A/H3N2*_3_, equaling the A/H3N2 incidence indicator for the period from Oct. 2018 to Jan. 2020, and equaling to 0 for other weeks. Similarly, we split the B/Yamagata incidence indicator into two, corresponding to the periods before and starting the 2017-2018 B/Yamagata epidemic. Finally, we note that it takes 1-2 weeks between influenza illness and influenza-associated mortality [1]. Correspondingly, we relate the mortality rate *M*(*t*) on week *t* to the *shifted* incidence indicators, e.g.

**Figure 2:**
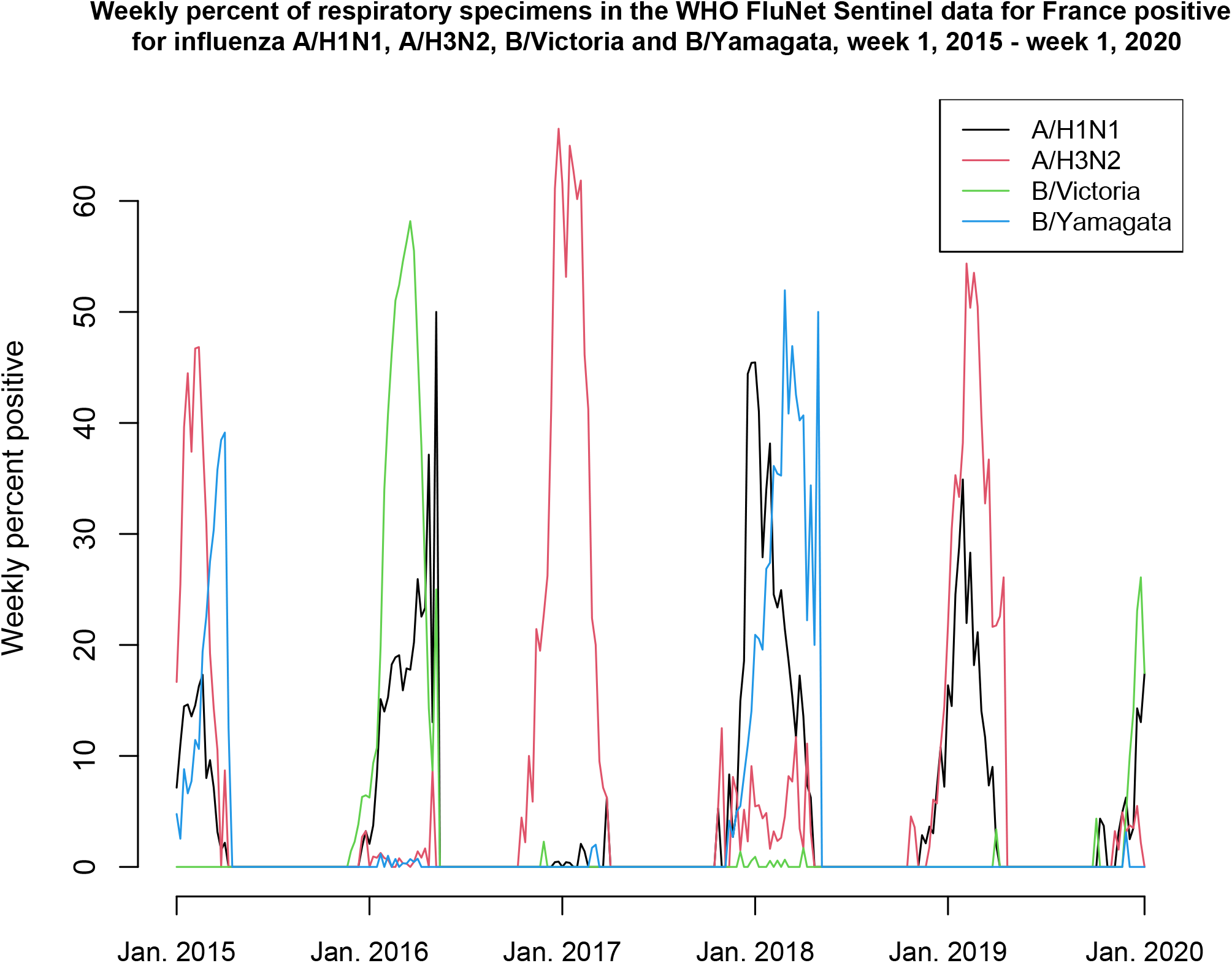
Weekly percent of respiratory specimens in the WHO FluNet Sentinel data for France positive for influenza A/H1N1, A/H3N2, B/Victoria and B/Yamagata, week 1, 2015 - week 1, 2020

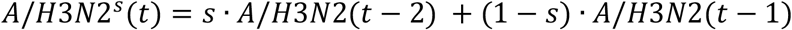

where the parameter *s* (common for all incidence indicators) is chosen to minimize the R-squared for the model fit. The regression model that we use is:

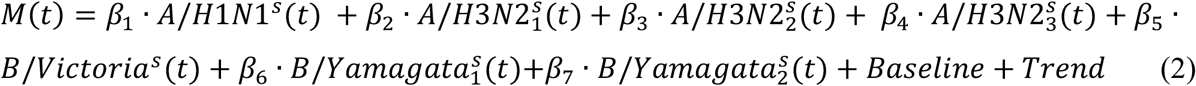

Here, Baseline represents weekly rates of mortality not associated with influenza circulation, and *Baseline*(*t*) is modelled to have annual periodicity in week *t*. We use periodic cubic splines to model the baseline mortality rates whose shape is unknown [1,6]. The trend *Trend*(*t*) is modelled as a quadratic polynomial in week *t*. Finally, to account for the autocorrelation in the noise, we use a bootstrap procedure (resampling the noise on different weeks) to estimate the confidence bounds for various quantities evaluated in the model [1].

## Results

Figure 1 plots the weekly mortality rates per 100,000 persons in France between week 3, 2015 – week 2, 2020, as well as the weekly rates of ILI consultations in metropolitan France during that period. While periods of excess mortality generally correspond to higher ILI levels (Figure 1), the relation between influenza circulation and excess mortality depends not only on ILI levels but also on the circulating influenza strains. Figure 2 plots the weekly percent of respiratory specimens in the WHO Flunet Sentinel data [20] that were positive for each of the major influenza subtypes: A/H1N1, A/H3N2, B/Victoria and B/Yamagata. Figure 2 shows that the majority of samples in the WHO FluNet sentinel data were positive for at least one influenza subtype during periods of high ILI levels. Figures 2 and 1 also show significant spikes in mortality during periods when influenza A/H3N2 and B/Yamagata circulation levels were high.

Figure 3 show the results of the fit for the model given by eq. 2. The model fit is generally temporally consistent except for the 2017-2018 season during which the circulating A/H1N1 and B/Yamagata strains had a different age distribution (Discussion). Figure 3 shows high levels of excess mortality associated with influenza A/H3N2 circulation during the 2016-2017 season, as well as the 2014-2015 season, and, to lesser extent, the 2018-2019 season. High levels of influenza-associated mortality corresponding to the circulation of influenza B/Yamagata and A/H1N1 took place during the 2017-2018 season. During the 2015-2016 season, the first peak in mortality was related to influenza circulation and was part of the baseline rates of mortality, with the second peak being associated with the circulation of influenza B/Victoria and A/H1N1.

**Figure 3:**
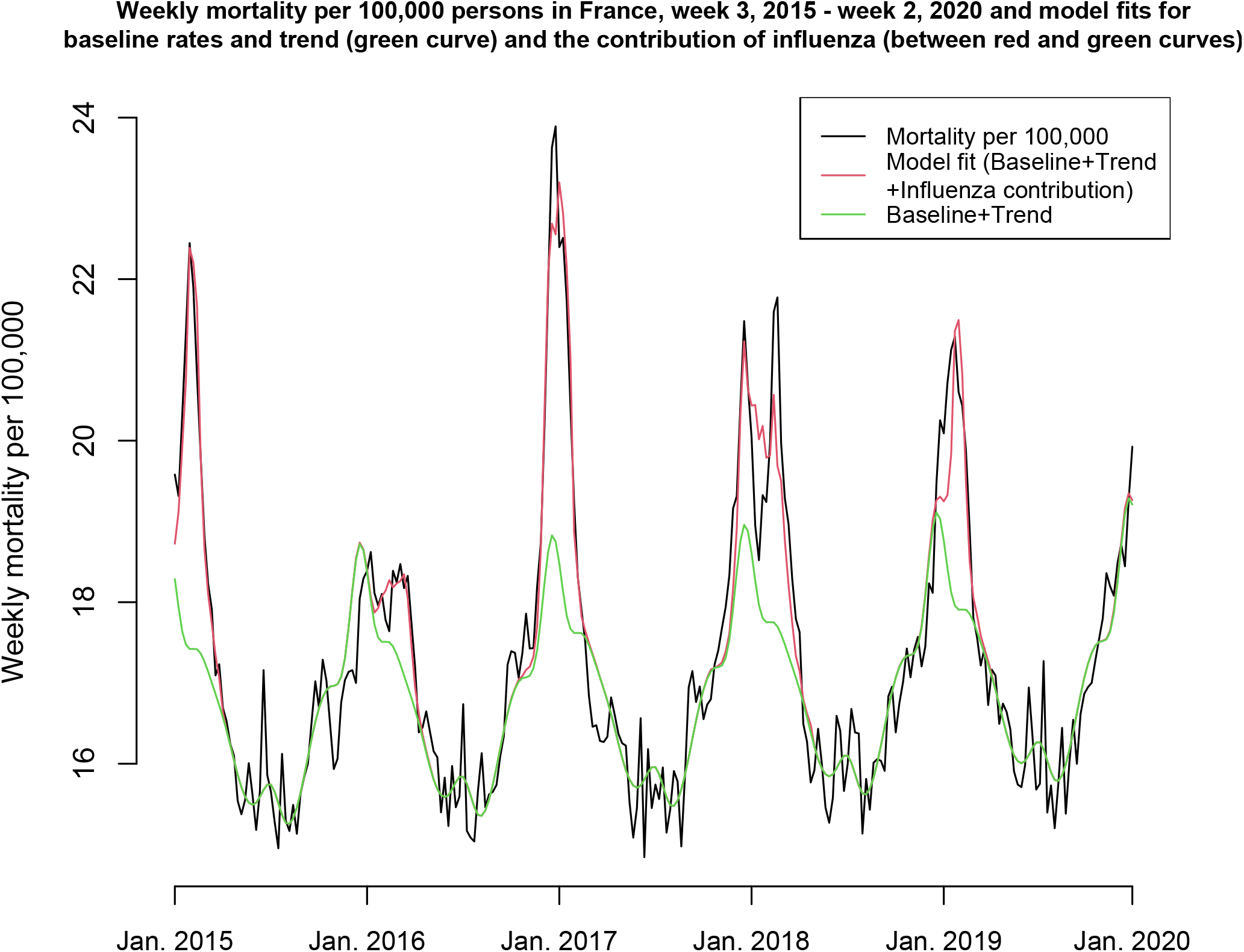
Weekly mortality per 100,000 persons in France, week 3, 2015-week 2, 2020 and model fits for baseline rates and trend (green curve) and the contribution of influenza (between red and green curves)

Table 1 suggests that during the study period, on average, there were 15403 (95% CI (12591,18229)) annual influenza-associated deaths, of which 60.3% (49.9%,71.9%) were associated with influenza A/H3N2, and 29.5% (13.3%,45.5%) were associated with influenza B/Yamagata. Additionally, during weeks when levels of ILI consultation in mainland France were above 50 per 100,000 persons [19], 7.9% (6.5%,9.4%) of all deaths in France were influenza-associated.

## Discussion

In the period preceding the COVID-19 pandemic, influenza epidemics resulted in high rates of influenza-associated mortality in different countries including the US [2], UK [3], and the EU countries [4,5]. Rates of excess mortality associated with influenza epidemics vary between countries (e.g. [13]), with some of the factors affecting that variability being influenza vaccination coverage levels in different populations groups and practices for testing for influenza and the use of antiviral medications during respiratory illness episodes. In France, high rates of excess mortality were found in the years preceding the COVID-19 pandemic [15]. In this paper we combined data on sentinel ILI surveillance [19] and sentinel virological surveillance [20] in France and used it in a previously developed regression model to estimate the number of deaths associated with the circulation of the major influenza subtypes (A/H3N2, A/H1N1, B/Yamagata and B/Victoria) in France between 2015-2019. We found that there were on average 15403 (95% CI (12591,18229)) annual influenza-associated deaths, of which 60.3% (49.9%,71.9%) were associated with influenza A/H3N2, and 29.5% (13.3%,45.5%) were associated with influenza B/Yamagata. During weeks when levels of ILI consultation in mainland France were above 50 per 100,000 persons, 7.9% (6.5%,9.4%) of all deaths in France were influenza-associated. Our results suggest that boosting influenza vaccination coverage in different population groups and testing for influenza in respiratory illness episodes during periods of active influenza (particularly influenza A/H3N2) circulation in combination with the use of antiviral medications is needed to mitigate the impact of influenza epidemics in France.

Influenza vaccination coverage levels in different population groups in France are moderate-to-low [16], and significantly lower compared to influenza vaccination coverage levels in the corresponding population groups in the US [17]. This pertains to both influenza vaccination coverage in risk groups for influenza-associated complications, as well as to population groups that play an important role in spreading influenza infections in the community. In particular, studies have found that children experience higher than average influenza infection rates [25] and play an important role in the spread of influenza in different countries [26], including France [27]. Another factor that has an effect on influenza-associated mortality is testing for influenza infection during respiratory illness episodes (including pneumonia hospitalizations) together with prompt use of antiviral medications [5]. Finally, influenza vaccine effectiveness in older individuals can be quite low, and types of influenza vaccines administered to older individuals play a role in preventing adverse outcomes associated with influenza infections. In the US, new ACIP recommendations stipulate that adults aged ≥65 years should preferentially receive any one of the following higher dose or adjuvanted influenza vaccines: quadrivalent high-dose inactivated influenza vaccine (HD-IIV4), quadrivalent recombinant influenza vaccine (RIV4), or quadrivalent adjuvanted inactivated influenza vaccine (aIIV4) [28].

Our results have some limitations. Influenza surveillance data in France pertains to mainland France [19], whereas we’ve used data on mortality for the whole of France. Additionally, sentinel data on testing for viral specimens [20] has a moderate sample size and may not represent all of France. We note that influenza epidemics exhibit a great deal of temporal synchrony [29,30] which should help address the above limitations. Finally, despite the fact that we split some of the influenza subtype incidence indicators into several time periods, where might still be temporal variability in the relation between the incidence indicators used in this paper and rates of associated mortality. For example, while model fits are generally temporally consistent (Figure 3), the model fit for the mortality data for the 2017-2018 season is worse compared to other influenza seasons, which might be related to the fact that the influenza subtypes that circulated during that season (A/H1N1 and B/Yamagata) have different age distributions feeding into one ILI data stream.

### Conclusions

Our results suggest high rates of influenza-associated mortality in the years preceding the COVID-19 pandemic in France, with around 60% of it being associated with the circulation of influenza A/H3N2. These results, in combination with data on influenza vaccination coverage in France [16] suggest that boosting influenza vaccination coverage in different population groups and testing for influenza in respiratory illness episodes (including pneumonia) during periods of active influenza (particularly influenza A/H3N2) circulation in combination with the use of antiviral medications is needed to mitigate the impact of influenza epidemics. Additionally, very high rates of influenza-associated mortality in individuals aged over 75y during certain influenza seasons in France [15] suggest that additional efforts are needed to better protect those populations during influenza epidemics, both in nursing homes and in the general community.

## Data Availability

This manuscript is based on aggregate, publicly available data that can be access through refs. 19,20, 22,23

https://www.sentiweb.fr/france/fr/?page=table

https://app.powerbi.com/view?r=eyJrIjoiNjViM2Y4NjktMjJmMC00Y2NjLWFmOWQtODQ0NjZkNWM1YzNmIiwidCI6ImY2MTBjMGI3LWJkMjQtNGIzOS04MTBiLTNkYzI4MGFmYjU5MCIsImMiOjh9

https://www.insee.fr/fr/statistiques/4931039?sommaire=4487854#tableau-figure1

https://www.insee.fr/fr/statistiques?debut=0&theme=1

